# SURGE CAPACITY OF CURATIVE SECTOR HEALTHCARE INSTITUTIONS FOR THE MANAGEMENT OF DISASTERS FOLLOWING DISEASE EPIDEMIC IN SRI LANKA

**DOI:** 10.1101/2023.09.19.23295791

**Authors:** R. M. Nayani Umesha Rajapaksha, Chrishantha Abeysena, Aindralal Balasuriya, Millawage Supun Dilara Wijesinghe, Suranga Manilgama

## Abstract

**Background:** The ability to obtain adequate staff, supplies, structures, and systems to provide sufficient care to meet the immediate needs of an influx of patients is defined as surge capacity. The research aimed to describe the surge capacity of curative-healthcare institutions for disaster management at disease endemic district of Sri Lanka.

**Methods:** From May to September 2019, a descriptive cross-sectional study was conducted among all curative-healthcare institutions (n=46) with inward-care facilities for patient care in the Kurunegala district, Sri Lanka. The ’Science of Surge Theory’ was used to develop an interviewer-administered tool. The medical administrator of the institution collected the data from a designated employee for disaster management in the institution. For the criteria of overall surge capacity for medical disaster management, the levels have been categorized as basic (26-50%), moderate (51-75%), and high (>75%).

**Results:** Response-rate was 93.5% (n=43). The higher proportion of the institutions had (55.8%; n=24), inadequate staff capacity, and lack of equipment. Of the equipped institutions, 13.3% (n=4), 38.7% (n=12), 40.9% (n=9), and 30.2% (n=13) had an adequate number of adjustable beds, infusion pumps, saturation monitors, and oxygen cylinders, respectively. The higher proportion of the institutions had designated emergency treatment units (90.7%; n=39). The least proportion of the institutions had X-ray services (11.6%, n=5), Ultrasound (USS) (9.3%; n=4) and blood bank services (9.3%; n=4). The higher proportion (69.8%; n=30) had inadequate overall surge capacity. The Provincial General Hospitals and 75% (n=3) of the Base-hospitals had moderate-level and higher proportion of the District Hospitals (76.3%; n=29) had basic level overall surge capacity for disaster management.

**Conclusion:** The overall surge capacity was inadequate and there is a need for improvement of surge capacity of the curative-healthcare institutions in the district for disease outbreak management. The capacity development programmes need to be initiated to improve the preparedness.

- **What is already known on this topic** Surge capacity refers to a healthcare system’s ability to respond to a sudden increase in patient care demands. Furthermore, surge capacity is a measurable representation of ability to manage a sudden influx of patients following emergencies and it depends on a well-functioning incident management system. The capacity assessments are more important than encouraging stakeholders for management during response. Therefore, capacity assessment and outbreak response planning should be aligned as a measure of outbreak preparedness.
- **What this study adds** The research aimed to describe the surge capacity of curative-healthcare institutions for disaster management following a disease epidemic in a disease-endemic major district of Sri Lanka. The overall surge capacity was inadequate and there is a need for improvement of surge capacity of the curative-healthcare institutions in the district for disease outbreak management. The capacity development programmes need to be initiated to improve the preparedness to face future epidemics/disasters.
- **How this study might affect research, practice or policy** The results of this study have many implications for practice, education, and research. With improving need of the rising population, there is increased risk of disasters including disease outbreaks. During such situations, healthcare institutions need to play a significant role. Assessing the surge capacity is the first step to obtaining baseline data about their capacity to respond to future disasters in their institutions. The findings of this study have determined critical areas of adequacy of staff, disaster planning and preparedness, adequacy of equipment, services, availability of guidelines, capabilities of risk communication to address the needs of healthcare providers in public sector institution for efficient and timely disaster response.

## Introduction

Surge capacity refers to a healthcare system’s ability to respond to a sudden increase in patient care demands [1]. Surge capacity, on the other hand, has three key dimensions. Surge capacity has three dimensions: ’healthcare facility-based surge capacity, public health surge capacity, and community-based surge capacity’ [2]. Healthcare facility- based surge capacity refers to a facility’s ability to treat an increased number of sick and injured patients, particularly when inpatient care is required [3]. Furthermore, surge capacity is required to respond effectively to events that result in a large influx of patients. The primary components of surge capacity are "staff",’’ stuff,’’ structure", and systems [4]. Therefore, the surge capacity is defined as ‘the ability to obtain adequate staff, supplies and equipment, structures and systems to provide sufficient care to meet immediate needs of an influx of patients following a large-scale incident or outbreak’ [5]. Personnel are referred to as staff, supplies and equipment are referred to as stuff, facilities and specific organisational structures such as Incident Command System (ICS) are referred to as structures, and systems include integrated management policies and processes [6,7,8]. The available quantity of them depends on the systems and processes. A system needs to establish to identify the medical needs, available resources, resource mobilization and manage them effectively to gain maximum outcome from the managerial process. Overall medical surge strategy refers to maximum utilization of available resources prior to resorting to the use of alternate care facilities following a disaster. However, the identification of staff, stuff and the structure that make healthcare delivery system and prioritize initial response activities incidents is a challenge, because healthcare workers are unfamiliar for their roles and less practical experience assessing mass casualty incidents [1, 6]. Furthermore, surge capacity is a measurable representation of ability to manage a sudden influx of patients following emergencies and it depends on a well-functioning incident management system [9]. Therefore, an assessment of surge capacity, including "staff, ’’stuff”, ’’structure”, and “systems” is a priority. Over the last few years, there has been an increase in the number of infectious disease outbreaks, and large-scale outbreaks affect the entire community [10,11]. If there is adequate public health surge capacity, the surge burden on clinical healthcare facilities will be reduced. As a result, emergency preparedness is more critical than emergency response [12]. Furthermore, the surge capacity should be conceptualized across all aspect of the healthcare delivery system prior to implement the policies. A system to coordinate and operationalize the process by balancing activity across all key areas and responding to escalating healthcare demands for surge capacity has to be developed to minimize the burden [6,13]. Importantly, the administrators of the healthcare institutions and authorized persons in each unit should be aware of their surge capacity for disaster management specially following outbreaks such as dengue, COVID-19 etc. Therefore, there is a clear need to assess the surge capacity of healthcare institutions to develop their surge capacity to respond for future events and develop the capacity development programs. Importantly, the ability to deliver optimal medical care in the setting of a mass casualty event, regardless of its cause, will in large part be dependent on an immediately available supply of key medical equipment, supplies, and pharmaceuticals [14]. Finally, the identified key components of surge capacity include the four S’s: ‘staff’, ‘stuff’, ‘structure’ and ‘systems’ [5].

When it is related to disease outbreaks, the epidemic of dengue fever is becoming more common and severe around globe, and it has become a major public health issue in Sri Lanka. Dengue fever has been on an upward trend in Sri Lanka for the past two decades. Dengue outbreaks severely impacted 15 of Sri Lanka’s 26 districts during the most recent major epidemic. In 2017, the Kurunegala district witnessed a significant increase in dengue cases. There is evidence that annual outbreaks have occurred in the Kurunegala district for the last two decades. Notably, the management capacity of the Kurunegala district’s government healthcare institutions was exceeded due to the dengue outbreak, leading to a disaster for the district in the recent outbreak. To achieve sustainable development in institutions, a systematic approach to capacity development programme planning is required. Before developing long-term capacity development strategies, capacity assessment surveys should be conducted to detect existing needs and demand for capacity development. However, identifying the items, personnel, systems, and structures that comprise the healthcare delivery system is challenging [15]. Therefore, there was a need to conduct a systematic assessment of the surge capacity of healthcare institutions including assessment of staff capacities, availability of disaster preparedness and response planning, nomination of the focal points and their experience and training disaster management, availability of equipment, services and ability of the emergency management team for risk communication, incidence commanding system, control activities during response phase, communication and coordination during an emergency, surge planning of staff, stuff, space, special activities, tracking of cases, triaging of cases, treatment for emergency case and transportation of the needy patients for special management procedures. Effective disaster management should be consistent with preparedness of outbreaks and anticipated response planning. However, it is neglected by disease endemic countries. The measures should be depended on the context of the countries. The capacity assessments are more important than encouraging stakeholders for management during response. Therefore, capacity assessment and outbreak response planning should be aligned as a measure of outbreak preparedness [16]. Furthermore, strengthening the capacity during preparedness phase to mitigate bad consequences during outbreak response is a responsibility of all the community members. Measures to ensure the effective use of resources including human resources, funds, and logistics for effective management of outbreak response activities need to be taken during the phase of the preparedness [17]. The aim of the study was to describe the surge capacity of the public sector curative healthcare institutions for the management of disasters in Kurunegala district, Sri Lanka.

## Methods

From June to October 2019, a descriptive cross-sectional study was carried out in the Kurunegala district of Sri Lanka. The public health system in Sri Lanka is divided into two major sectors: curative and preventive. The curative care health sector consists of 46 healthcare institutions that provide inpatient care for patients. The survey included one Provincial General Hospital (PGH), one "A" grade Base Hospital (BH), three "B" grade BH, nine "A" grade District Hospitals (DH), 11 "B" grade DH, and 21 "C" grade DH. The study population was accessed using an Interviewer Administered Questionnaire (IAQ). The IAQ was formulated according to ‘the Science of Surge Theory’ for surge capacity assessment of the healthcare institutions after comprehensive exploring the litreture on concept of surge capacity [1,5,18]. Seven consensus statements were generated for evaluation of ‘The Science of Surge’, which emphasize the importance of funded research in the area of surge capacity metrics; the utility of research registry; the purpose of the data for policy planning and administration; the importance of data management system; and the basic necessity of a criterion standard metric for quantifying surge capacity [18]. The service of a panel of experts from different professional backgrounds, some of them have been directly involved in the managing disease outbreaks in curative sector (Consultant Physicians), Consultant Community Physicians, Consultant Epidemiologist, Consultants in Medical administration as well as the qualified disaster managers in the country. Two qualified medical graduates who have Master of Science in Community Medicine and Medical Administration were trained as data collectors. The data collectors were trained by the principal investigator on how to collect the data and how to enter on the data sheet including obtaining informed written consent, ensuring confidentiality, administering the questionnaire and on uniform administration of every question. At the end of the training session, the data collectors were given the opportunity to conduct mock interviews and these interviews were observed by the principal investigator and constructive feedbacks were given. Pre-testing was conducted in similar settings in Gampaha district to identify the feasibility issues. Data was collected by the principal investigator and trained two medical officers by conducting interviews. The authorized officer as the focal point for disaster management of healthcare institutions in the Kurunegala district by the head of the institution were invited to assess the surge capacity of their respective healthcare institutions. An IAQ was administered by the trained interviewers in a separate place at a free time without disturbance for their routine work.

### Staff and Cadre Approval for the Institutions

The available total staff, the healthcare staff who directly involving outbreak management and their cadre for all institutions were taken by the data collectors. The cadre positions of the healthcare workers and supportive workers were extracted from the official updated document of Ministry of Health in 2017 to compare with available staff to decide the adequacy of the staff. Infrastructure capacity, annual service provision and population drain for curative sector institutions were considered for determining the cadre provision by the Ministry of Health Sri Lanka. Equal or more than the 75% of the required cadre positions was considered as ‘adequate number of staff’ for the institutions after taking expert opinion. The availability of a focal point and his/her experience and training were assessed during the interview. The availability of the disaster preparedness and response plan, availability of the equipment, outbreak management guidelines were observed. Because of unavailability of the proper equipment management system in the institutions, the perceived level of adequacy of the available equipment by the authorized person of the institution was compared to the monthly consumption of the items during 2017 dengue outbreak. The average monthly requirement of equipment during the last disease outbreak season (Dengue epidemic) were compared with the currently available monthly stock. If currently available monthly stock is similar or more than 75% of the average consumption during the outbreak season, it was considered as “adequate consumable in place”.

### Overall Surge capacity

An aggregated percentage mark of each category of the IAQ was analyzed and described according to a ranking system considering four levels of capacity components including ‘Clear need for increased capacity (≤25%), Basic level of capacity in place (26 – 50%), Moderate level of capacity in place (51 – 75%) and High level of capacity in place (>75%). The range was zero to 100%. The mean completion time of the tool was found to be approximately 30 minutes.

## Results

### KEY COMPONENT ONE: STAFF

The median cadre of the curative sector staff per institution was 49 (IQR 27 - 105). The median available number of curative sector staff per institution was 42 (IQR 21 - 74). The higher proportion of the curative sector healthcare institution had inadequate staff capacity (55.8%; n=24) (Table 1).

**Table 1.**
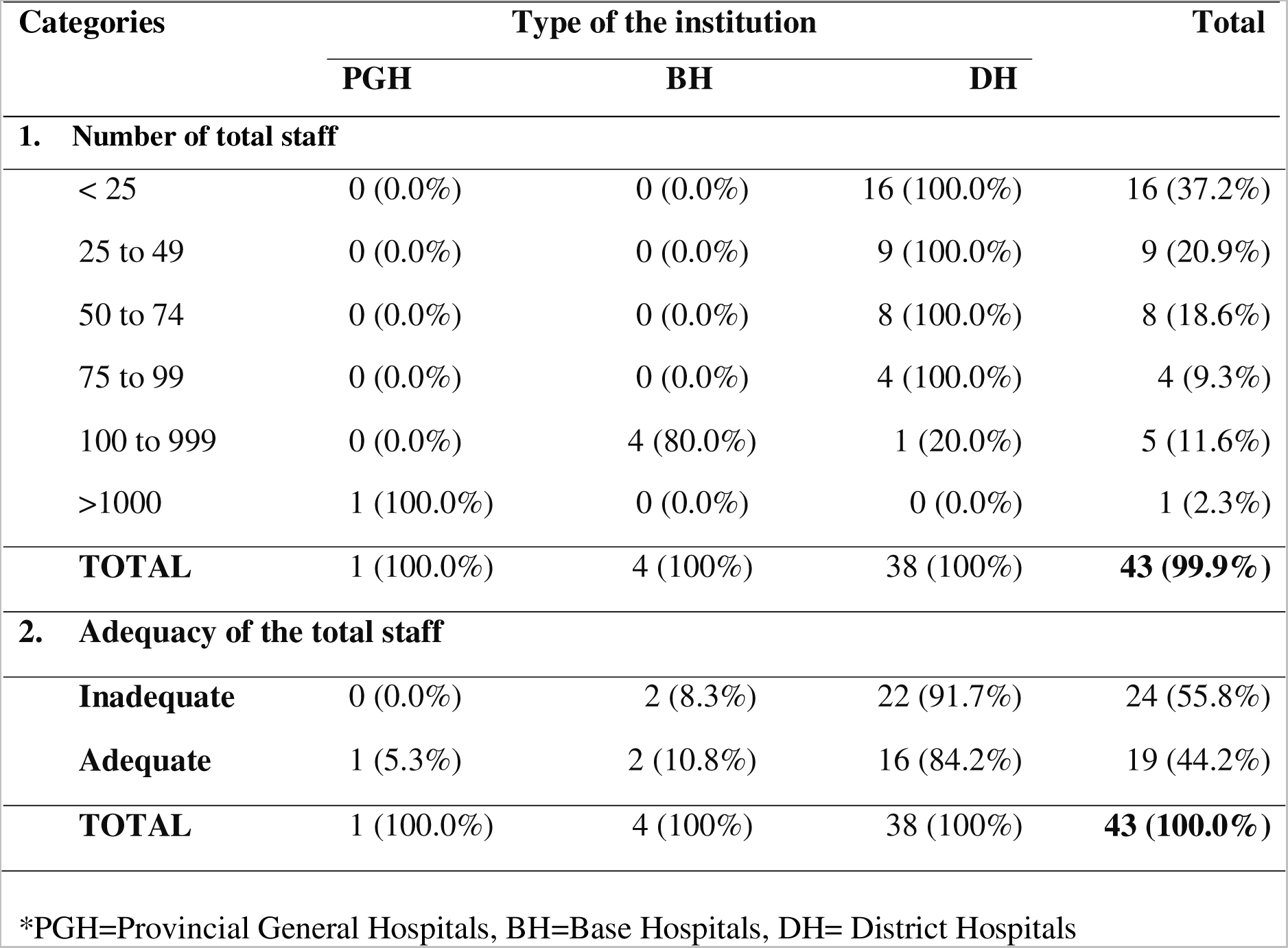
Number of Staff in The Different Type of the Institutions.

### Medical Team

The cadre of specialist medical officers was 102 for the district. Ranging from one to 74. There were 95 specialist medical officers who have been working at six healthcare institutions in the district. Of them, higher proportion of the institutions (50%; n=3) had five to nine specialist medical officers and 50% (n=3) of the institutions had adequate specialist medical officer capacity. All curative sector institutions had medical officers (100.0%; n=43). The cadre of medical officers was 803 for the 43 curative sector institutions in the district range from one to 416. There were 733 medical officers working in the district. Of them, higher proportion of the institutions (39.5%; n=17) had three to five medical officers and 76.7% (n=33) of the institutions had adequate medical officer capacity (Table 2). The cadre of nursing officers was 2423 for the district ranging from one to 1269. There were 2133 nursing officers working in 31 curative sector healthcare institutions in the district. Of them, higher proportion of the institutions (58.1%; n=18) had one to 25 nursing officers and 51.2% (n=22) of the institutions had adequate nursing officer capacity (Refer Table S1).

**Table 2.** Adequacy of the Healthcare Medical Staff for the Curative Sector Institutions for Disaster Management.

| Medical team |  | Type of the institution |  |  | Total |
| --- | --- | --- | --- | --- | --- |
|  |  | PGH | BH | DH |  |
| **1** | **Specialists MOO** |  |  |  |  |
|  | Adequate | 1 (100.0) | 2 (40.0%) | NA | **3 (50.0%)** |
|  | Inadequate | 0 (0.0%) | 3 (60.0%) | NA | **3 (50.0%)** |
|  | **Total** | **1 (100%)** | **5 (100%)** | **NA** | **6 (100.0%)** |
| **2** | **MOO** |  |  |  |  |
|  | Adequate | 1 (100%) | 1 (25.0%) | 31 (81.6%) | **33 (76.7%)** |
|  | Inadequate | 0 (0.0%) | 3 (75.0%) | 7 (18.4%) | **10 (23.3%)** |
|  | **Total** | **1 (100%)** | **4 (100%)** | **38 (100%)** | **43 (100.0%)** |
| **1.** | **Nursing staff** |  |  |  |  |
|  | Adequate | 1 (100.0) | 2 (50.0%) | 19 (50.0%) | **22 (51.2%)** |
|  | Inadequate | 0 (0.0%) | 2 (50.0%) | 19 (50.0%) | **21 (48.8%)** |
|  | **Total** | **1 (100%)** | **4 (100%)** | **38 (100%)** | **43 (100.0%)** |

\*MOO=Medical Officers; NA=Not applicable
\*PGH=Provincial General Hospitals, BH=Base Hospitals, DH= District Hospitals

### Paramedical team

The cadre of PHI was two for the curative sector institutions of the district and three PHII were available. Out of the 43 institutions, 7% (n=3) did not have cadre provisions for PHFO. The cadre of PHFO was 39 for the curative sector institutions of the district and five PHFO were available. Of the institutions which have cadre provisions, only 12.5% (n=5) had adequate PHFO capacity. Out of the 43 institutions, 25.6% (n=11) did not have midwives. The cadre of midwives was 317 for the curative sector health institutions of the district. There were 172 midwives working in 32 institutions in the district. Of them, higher proportion of the institutions (34.9%; n=15) had one to two midwives and only 9.3% (n=4) of all institutions had adequate midwives’ capacity. All curative sector institutions have Pharmacy staff including pharmacists and/or dispenser (100.0%; n=43). The cadre of pharmacy staff was 197 for the district, range from one to 50. The available number was 174 in all 43 institutions in the district. The higher proportion of the institutions (51.2%; n=22) had two pharmacy staff members and 74.4% (n=32) of the institutions had adequate pharmacy staff capacity (Table 3). Laboratory technicians are Medical Laboratory Technicians (MLTs) or Public Health Laboratory Technicians (PHLTs). Out of the 43 institutions, 46.5% (n=20) did not have laboratory technicians and 34.9% (n=15) did not have an approved cadre by 2019. The carder of laboratory technicians was 140 for the district range from one to 42 and. There were 95 technicians available in the district. Out of the available institutions, least proportion of the institutions (21.7%; n=5) had more than three laboratory technicians. Out of the institution which has approved carder positions (n=28), 42.9% (n=12) of the institutions had adequate laboratory technician’s capacity (Refer Table S2).

**Table 3.** Adequacy of Curative Sector Para-medical Staff for Disaster Management.

| Para medical team |  | Type of the institution |  |  | Total |
| --- | --- | --- | --- | --- | --- |
|  |  | PGH | BH | DH |  |
| **1** | **PHII** |  |  |  |  |
|  | Adequate | 1 (100.0) | 1 (100.0) | NA | **2 (100.0%)** |
|  | Inadequate | 0 (0.0%) | 0 (0.0%) | NA | **0 (0.0%)** |
|  | **Total** | **1 (100%)** | **1 (100%)** |  | **2 (100.0%)** |
| **2** | **PHFO** |  |  |  |  |
|  | Adequate | NA | 1 (33.3%) | 34 (91.9%) | **35 (87.5%)** |
|  | Inadequate | NA | 2 (66.7%) | 3 (8.1%) | **5 (12.5%)** |
|  | **Total** |  | **3 (100%)** | **37 (100%)** | **40 (100.0%)** |
| **3** | **Midwives** |  |  |  |  |
|  | Adequate | 0 (0.0%) | 1 (25.0%) | 3 (7.9%) | **4 (9.3%)** |
|  | Inadequate | 1 (100.0) | 3 (75.0%) | 35 (92.1%) | **39 (90.7%)** |
|  | **Total** | **1 (100%)** | **4 (100%)** | **38 (100%)** | **43 (100.0%)** |
| **4** | **Pharmacy staff** |  |  |  |  |
|  | Adequate | 1 (100.0) | 4 (100.0%) | 27 (71.1%) | **32 (74.4%)** |
|  | Inadequate | 0 (0.0%) | 0 (0.0%) | 11 (28.9%) | **11 (25.6%)** |
|  | **Total** | **1 (100%)** | **4 (100%)** | **38 (100%)** | **43 (100.0%)** |
| **5** | **Lab Technicians** |  |  |  |  |
|  | Adequate | 1 (100.0) | 2 (50.0%) | 9 (39.1%) | **12 (42.9%)** |
|  | Inadequate | 0 (0.0%) | 2 (50.0%) | 14 (60.9%) | **16 (57.1%)** |
|  | **Total** | **1 (100%)** | **4 (100%)** | **23 (100%)** | **28 (100.0%)** |

\* PHII= Public Health Inspectors, PHFO=Public Health Field Officers, PGH=Provincial General Hospitals, BH=Base Hospitals, DH= District Hospitals

### Supportive staff

All curative sector institutions have health care assistants (100.0%; n=43). The cadre of health care assistants was 2370 for the district range from seven to 1082. There available number was 2118. The higher proportion of the institutions (34.9%; n=15) had 21 to 40 health care assistants and 79.1% (n=34) of the institutions had adequate health care assistant’s capacity (Table 4). All curative sector institutions have drivers (100.0%; n=43). The cadre of drivers was 73 for the district range from one to 16. There available number was 67. Of them, higher proportion of the institutions (83.7%; n=36) had only one driver and 97.7% (n=42) of the institutions had adequate drivers’ capacity. Out of the 43 institutions, one institution did not have a cadre provision and 16.3% (n=7) did not have management assistants. The cadre of management assistants was 150 for the curative sector health care institutions of the district. There were 107 management assistants working in 36 curative sector health care institutions in the district. Of them, higher proportion of the institutions (44.2%; n=18) had one management assistants and 42.9% (n=18) of the institutions had adequate management assistants’ capacity (Refer Table S3).

**Table 4.** Adequacy of the Healthcare Supportive Staff for the Curative Sector Institutions for Disaster Management.

| Supportive staff |  | Type of the institution |  |  | Total |
| --- | --- | --- | --- | --- | --- |
|  |  | PGH | BH | DH |  |
| **1** | **Healthcare assistants** |  |  |  |  |
|  | Adequate | 0 (0.0%) | 1 (25.0%) | 3 (7.9%) | **4 (9.3%)** |
|  | Inadequate | 1 (100.0) | 3 (75.0%) | 35 (92.1%) | **39 (90.7%)** |
|  | **Total** | **1 (100%)** | **4 (100%)** | **38 (100%)** | **43 (100.0%)** |
| **2** | **Drivers** |  |  |  |  |
|  | Adequate | 0 (0.0%) | 4 (100.0%) | 38 (100.0%) | **42 (97.7%)** |
|  | Inadequate | 1 (100.0%) | 0 (0.0%) | 0 (0.0%) | **1 (2.3%)** |
|  | **Total** | **1 (100%)** | **4 (100%)** | **38 (100%)** | **43 (100.0%)** |
| **3** | **Management Assistants** |  |  |  |  |
|  | Adequate | 1 (100.0) | 3 (75.0%) | 14 (37.8%) | **18 (42.9%)** |
|  | Inadequate | 0 (0.0%) | 1 (25.0%) | 23 (62.2%) | **24 (57.1%)** |
|  | **Total** | **1 (100%)** | **4 (100%)** | **37 (100%)** | **43 (100%)** |

\*PGH=Provincial General Hospitals, BH=Base Hospitals, DH= District Hospitals

### KEY COMPONENT TWO: SUPPLIES FOR THE CURATIVE SECTOR

The higher proportion the institutions had adjustable beds (69.8%; n=30), infusion pumps (72.1%; n=31), and saturation monitors (51.2%, n=22) for dengue management activities. Oxygen cylinders were available in all the institutions (100%; n=43) and 27.9% (n=12) had Pack Cell Volume (PCV) monitors. Of the equipped institutions, 13.3% (n=4) had adequate number of adjustable beds, 38.7% (n=12) had number of infusion pumps, 40.9% (n=9) had adequate number of saturation monitors, 30.2% (n=13) had adequate number of oxygen cylinders and higher proportion (66.7%; n=8) had adequate PCV monitors (Table 5).

**Table 5.**
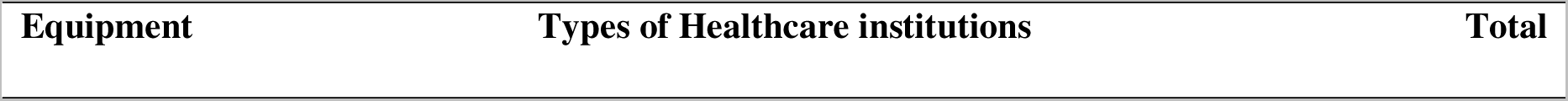
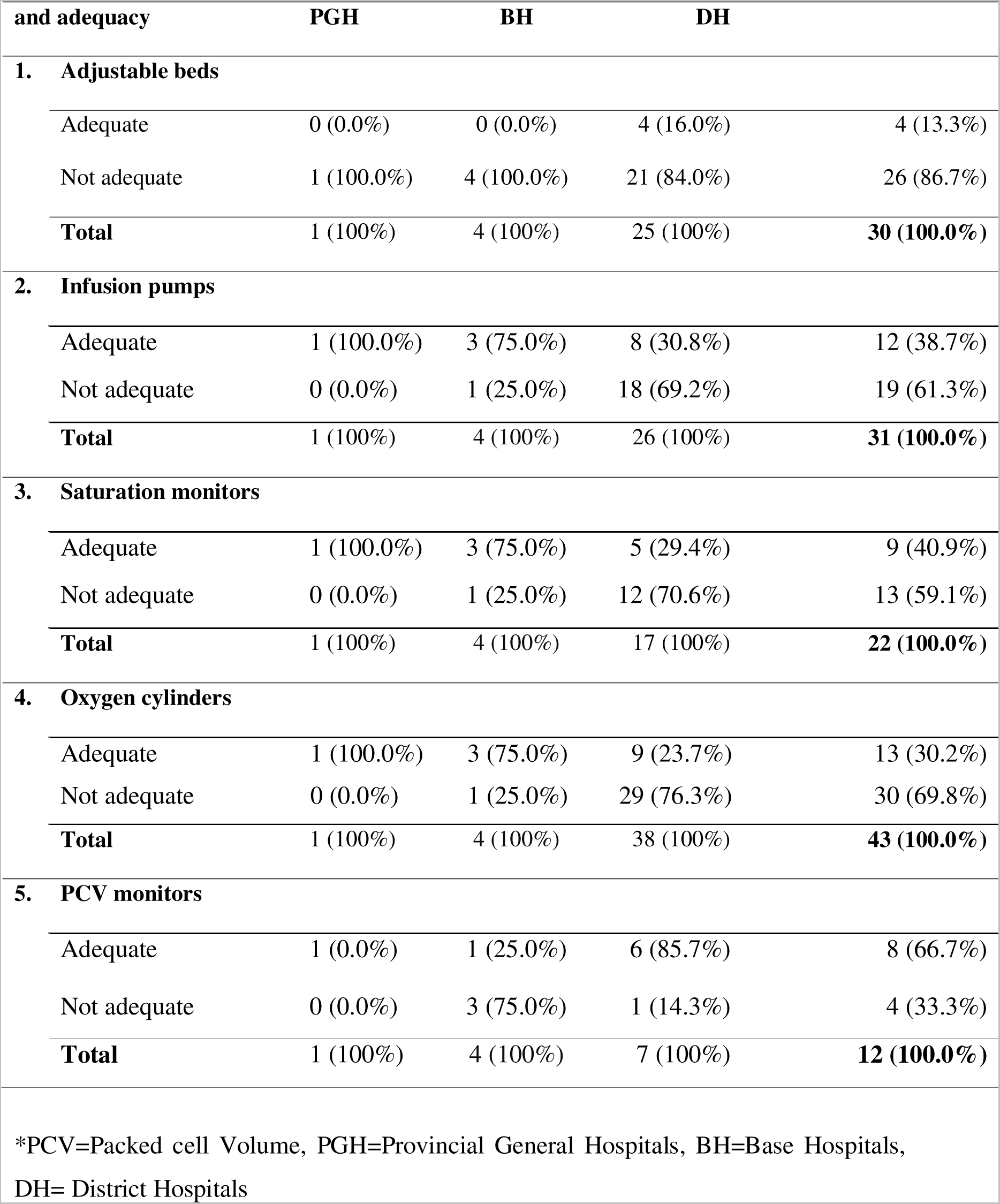
Adequacy of Equipment for Disaster Management in the Curative Sector Institutions.

| Equipment | Types of Healthcare institutions |  |  | Total |
| --- | --- | --- | --- | --- |

| and adequacy | PGH | BH | DH |  |
| --- | --- | --- | --- | --- |
| **1. Adjustable beds** |  |  |  |  |
| Adequate | 0 (0.0%) | 0 (0.0%) | 4 (16.0%) | 4 (13.3%) |
| Not adequate | 1 (100.0%) | 4 (100.0%) | 21 (84.0%) | 26 (86.7%) |
| **Total** | **1 (100%)** | **4 (100%)** | **25 (100%)** | **30 (100.0%)** |
| **2. Infusion pumps** |  |  |  |  |
| Adequate | 1 (100.0%) | 3 (75.0%) | 8 (30.8%) | 12 (38.7%) |
| Not adequate | 0 (0.0%) | 1 (25.0%) | 18 (69.2%) | 19 (61.3%) |
| **Total** | **1 (100%)** | **4 (100%)** | **26 (100%)** | **31 (100.0%)** |
| **3. Saturation monitors** |  |  |  |  |
| Adequate | 1 (100.0%) | 3 (75.0%) | 5 (29.4%) | 9 (40.9%) |
| Not adequate | 0 (0.0%) | 1 (25.0%) | 12 (70.6%) | 13 (59.1%) |
| **Total** | **1 (100%)** | **4 (100%)** | **17 (100%)** | **22 (100.0%)** |
| **4. Oxygen cylinders** |  |  |  |  |
| Adequate | 1 (100.0%) | 3 (75.0%) | 9 (23.7%) | 13 (30.2%) |
| Not adequate | 0 (0.0%) | 1 (25.0%) | 29 (76.3%) | 30 (69.8%) |
| **Total** | **1 (100%)** | **4 (100%)** | **38 (100%)** | **43 (100.0%)** |
| **5. PCV monitors** |  |  |  |  |
| Adequate | 1 (0.0%) | 1 (25.0%) | 6 (85.7%) | 8 (66.7%) |
| Not adequate | 0 (0.0%) | 3 (75.0%) | 1 (14.3%) | 4 (33.3%) |
| **Total** | **1 (100%)** | **4 (100%)** | **7 (100%)** | **12 (100.0%)** |

\*PCV=Packed cell Volume, PGH=Provincial General Hospitals, BH=Base Hospitals, DH= District Hospitals

### KEY COMPONENT THREE: STRUCTURE FOR DISASTER MANAGEMENT

The higher proportion of the institutions had total bed strength of one to 25 beds per institution (41.9%; n=18) and designated emergency treatment units (90.7%; n=39) (Refer Supplementary Table 4). Only 4.7% (n=2) had HDU and 7% (n=3) had Medical ICU facilities. The all-curative sector institutions with technicians have provided X-ray services (100%, n=5), Ultrasound service (USS) (100%; n=4) and blood bank services (100%; n=4). The higher proportion (62.8%; n=27) of the health care institutions had public health units, ECG services (83.3%; n=35) and medical laboratory services (51.2%; n=22) (Refer Table S5). Out of the available laboratory services (n=22), the higher proportion (59.1%; n=13) had taken average four to six hours to issue FBC reports (Refer Table S6).

### Overall surge capacity

There is one Provincial General Hospitals (PGH) in the district which has moderate level overall surge capacity for outbreak management. Out of the four base hospitals, higher proportion (75%; n=3) had moderate level surge capacity for outbreak management. Out of 38 District Hospitals (DH), higher proportion (76.3%; n=29) had basic level surge capacity for outbreak management (Table 6).

**Table 6.**
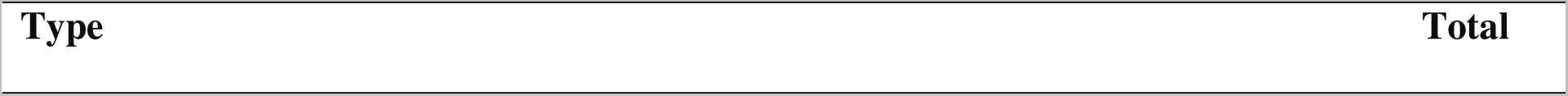
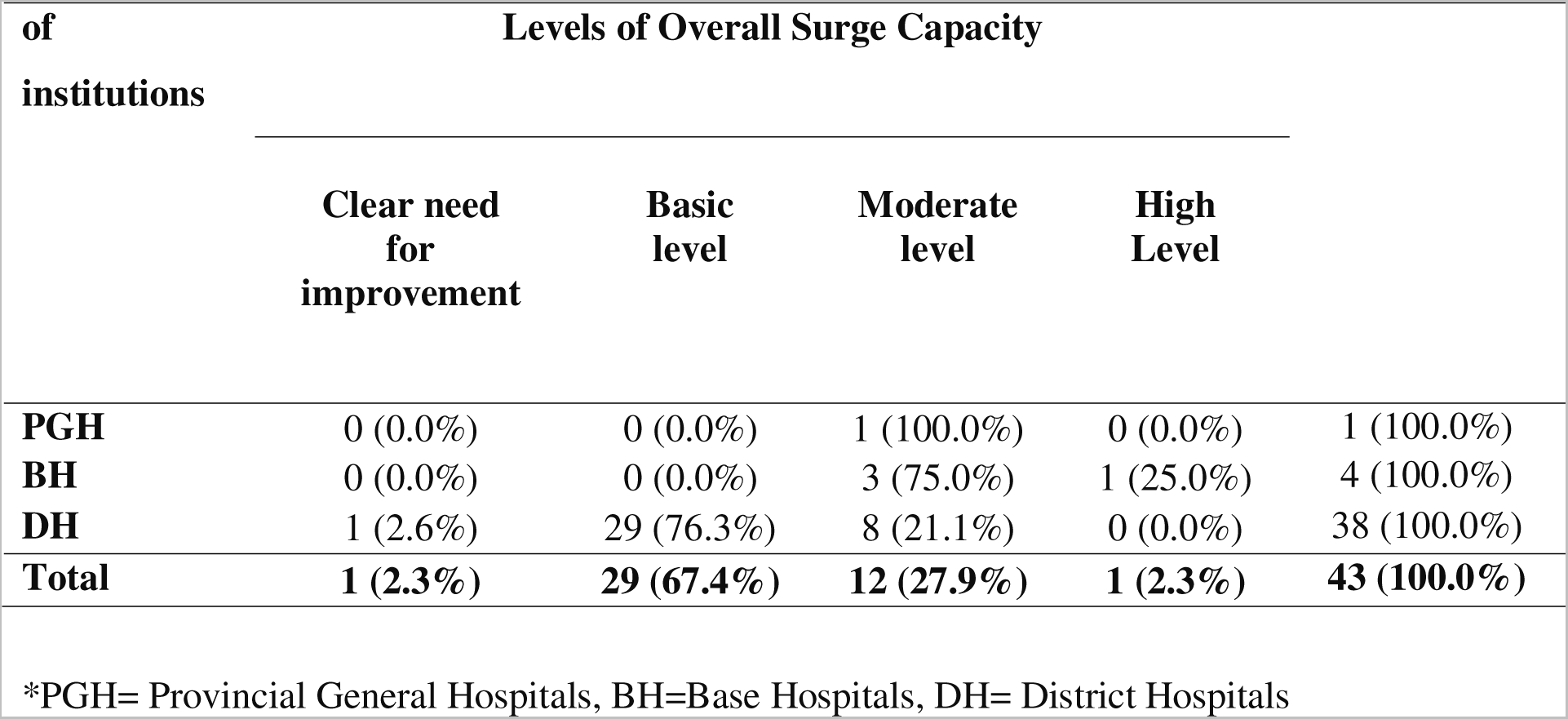
Overall Surge Capacity of the Public Sector Curative Healthcare Institutions in Kurunegala District for the Management of Outbreaks in 2019.

| of<br>institutions | Levels of Overall Surge Capacity |  |  |  |  |
| --- | --- | --- | --- | --- | --- |
|  | Clear need<br>for<br>improvement | Basic<br>level | Moderate<br>level | High<br>Level |  |
| **PGH** | 0 (0.0%) | 0 (0.0%) | 1 (100.0%) | 0 (0.0%) | 1 (100.0%) |
| **BH** | 0 (0.0%) | 0 (0.0%) | 3 (75.0%) | 1 (25.0%) | 4 (100.0%) |
| **DH** | 1 (2.6%) | 29 (76.3%) | 8 (21.1%) | 0 (0.0%) | 38 (100.0%) |
| **Total** | **1 (2.3%)** | **29 (67.4%)** | **12 (27.9%)** | **1 (2.3%)** | **43 (100.0%)** |

\*PGH= Provincial General Hospitals, BH=Base Hospitals, DH= District Hospitals

## Discussion

The higher proportion of the curative healthcare institutions (55.8%) had inadequate total staff capacity. Of them, the majority of institutions had adequate capacity of the MOO (76.7%), nursing staff (51.2%), pharmacy staff (74.4%), drivers (97.7%), PHII (100%), healthcare assistants (79.1%), and inadequate capacity of laboratory technicians (57.1%), PHMM (90.7%) and management officers (57.1%). The higher proportion the institutions had adjustable beds (69.8%), infusion pumps (72.1%), and saturation monitors (51.2%) for dengue management activities. Oxygen cylinders were available in all the institutions and only one-fourth (27.9%) had PCV monitors. Of the equipped institutions, only 13.3% had adequate number of adjustable beds, 38.7% had number of infusion pumps, 40.9% had adequate number of saturation monitors, 30.2% had adequate number of oxygen cylinders and majority (66.7%) had adequate PCV monitors. There were adequate public health services, ECG services, and medical laboratory services in the curative healthcare institutions. The majority of them have allocated separate units as Emergency Departments (ED). However, the bed strength was inadequate in the majority of the healthcare institutions. Moreover, bed strength in ED also inadequate. The availability treatment guidelines for adults were adequate. However, the availability of the other treatment guidelines and preventive guidelines were inadequate. Three-fourth of the curative healthcare institutions had nominated focal points for dengue management. Of them, 42.4% had experience in managing medical disasters, 31.6% had experience in managing both types of disasters and only one-fourth had undergone formal training on disaster management during their lifetime. Out of the 43 institutions, only 6.5% (n=2) completed the medical disaster management. Three-fourth (72.1%) of the institutions had written plans and 21.2% of them have evaluated their plans by conducting drills or simulations. Out of the preventive sector healthcare institutions, the higher proportion had inadequate surge capacity for the management of dengue. Of the curative healthcare institutions in the district, the PGH had moderate level overall surge capacity for dengue outbreak management. Out of the four BH, one-fourth had high level and three-fourth had moderate level surge capacity for outbreak management. Out of 38 DH, three fourth had basic level surge capacity for outbreak management. Since assessment of surge capacity involves assessment of managerial information, it is possible that the administrator or authorized person would not reveal true information. Thus, before collecting data the institution was given the information sheet which elaborated in detail the measures taken to ensure confidentiality and further during the training of data collectors special emphasis was given on developing a good rapport with the participants to minimize respondent variation and ensured that the individual data would not publish anywhere. The pre- testing of the tool was conducted to check acceptability, availability of such information in similar settings at Gampaha district. Single entry with random checking was done as opposed to double entry to improve the data quality.

### A) KEY COMPONENT ONE: Staff for emergency management

The present study revealed that the majority of the public sector healthcare institutions of the district had inadequate total healthcare staff capacity. However, 69.2% had adequate medical officers’ capacity in the preventive healthcare institutions and 76.7% of the curative healthcare institutions had adequate medical officers’ capacity. According to a study in China, there was a shortage of all categories of healthcare workers for emergency response and the medical officers who are responsible for community health work showed significantly higher response capacity than other healthcare staff [19]. Provision of urgent care during disasters, assist in policy making relevant to disaster preparedness, and become educated in disaster preparedness and management are obligations of medical officers [20]. Medical officers’ leadership, proactive anticipating and preparing for disasters is vital for increasing the effectiveness of the health system’s large-scale crisis response efforts [21]. Therefore, the adequate availability of medical officers for the healthcare institutions may strengthen the surge capacity of the public health sector institutions. In the present study, the higher proportion of the preventive healthcare institution had 30 to 45 staff members per institution and 69.2% had inadequate total staff capacity for outbreak prevention. Of them, 69.2% had inadequate PHII capacity, 80.8% had inadequate number of spray machine operators and entomology assistants for the dengue prevention activities. According to the study in Thailand, a division for dengue control usually consists of one district health officer, the number of other healthcare workers varies by district between two and 16, and one or two other supportive staff [22]. Available staff is an asset for the nation because they are the one who work as frontline workers during any outbreak situation in the country. Without the contribution of preventive sector staff, whole curative sector has to face critical challenging situation because of limited resources.

### B) KEY COMPONENT TWO: Stuff

The present study revealed that the majority of the preventive healthcare institutions had Personal Protective Equipment (PPE) (65.4%), chemical for mosquito control (92.3%), and fogging instrument (88.5%) for dengue management activities. Of them, 41.2% had adequate amount of PPE, 8.3% had adequate amount of chemicals and 17.4% had adequate fogging instruments. According to a study among district healthcare workers in Bangkok, Thailand, there is a lack of protective equipment such as masks, protective overalls after the fumigation visit following dengue prevention activities [22]. More than 50% of the respondents of the institutions of the present study perceived that they have inadequate supply of PPEs which leads to low level of surge capacity. In the present study, 73.1% of the preventive sector institutions did not have establish mechanism with instruments for waste management. Of the available institutions with proper waste management, one-fourth (28.6%) had adequate waste management equipment to prevent vector borne diseases. Dengue is a vector borne disease and waste management is the mainstay of dengue prevention. If there is no proper mechanism in the institutions, they cannot promote that good habit among the community. That may invariably lead to lack of waste management among the community too. This study revealed that the majority of the curative care institutions had one or more adjustable beds (69.8%), infusion pumps (72.1%), and saturation monitors (51.2%) for dengue patient management. Oxygen cylinders were available in all the institutions and 27.9% of them had PCV machines for monitoring of cases. Of the equipped institutions, 13.3% had adequate number of adjustable beds, 38.7% had adequate number of infusion pumps, 40.9% had adequate number of saturation monitors, 30.2% had adequate number of oxygen cylinders and 66.7% had adequate PCV machines. According to a case study among twelve public sector healthcare institutions situated in different regions of Sri Lanka, around 42% of healthcare equipment is not functional and therefore unusable [23]. The situation of the Sri Lanka is little better than Yemen and Ghana but worse than Costa Rica. However, in developed countries this performance figure is very high and, in some cases, close to 100% [24]. Administrators or coordinators may need to plan for means to supply alternate-care sites with oxygen, point-of-care testing equipment, walkers, wheelchairs, personal protective equipment, and other supplies [25,26]. Healthcare workers need to allocate beds, ventilators, and other supplies in a manner consistent with the goal of saving the most lives following a disaster [27,28]. Medical equipment management is important as human resource management to function a hospital in a maximum capacity. According to a study in District Base Hospitals, Kalutara District in Sri Lanka, all hospitals reported lack of efficient equipment management system and all hospitals were found to be without guidelines for procurement of equipment. Of all departments in the healthcare institutions, 85.4% did not have proper register maintained for equipment maintenance [29]. The present study also revealed that there is no equipment management system in the majority of the healthcare institutions. Therefore, the adequacy of the equipment was assessed subjectively. If there is an efficient equipment management system, it would have done objectively which may increase the quality of collected data. Therefore, all the disaster coordinators or focal points should plan for impending disasters and all healthcare institutions should be equipped with adequate stocks of equipment which leads to development of the surge capacity of the relevant setting. Although Sri Lanka has experienced different forms of disasters, the administrative, and health were simply not able to respond rapidly to the workload demands created by the disaster because of inadequate surge capacity [30]. A capacity assessment is needed to perform to identify the activities required to carry out on a priority basis to overcome the temporary mismatch between demand and supply of care following disasters, in view of developing the hospital preparedness capacity to respond effectively [31]. There is a worldwide urgent need of capacity building among health care providers in disaster management in order to achieve desired goals of management of disasters [32]. Although a comprehensive literature search was conducted, the investigators could not find research on the surge capacity assessment for the management disease outbreaks among healthcare institutions in Sri Lanka before COVID-19 outbreak. The Incident Command System (ICS) for out break management was separately published [33].

## Conclusion

The overall surge capacity was inadequate and there is a need for improvement of surge capacity of the curative-healthcare institutions in the district for disease outbreak management. The capacity development programmes need to be initiated to improve the preparedness.

### Implications

The results of this study have many implications for practice, education, and research. With improving need of the rising population, there is increased risk of disasters including disease outbreaks. During such situations, healthcare institutions need to play a significant role. Assessing the surge capacity is the first step to obtaining baseline data about their capacity to respond to future disasters in their institutions. Effective disaster planning, training and education initiatives rely on inputs from the target population before designing the goals and objectives for such initiatives. The findings of this study have determined critical areas of adequacy of staff, disaster planning and preparedness, adequacy of equipment, services, availability of guidelines, capabilities of risk communication to address the needs of healthcare providers in public sector institution for efficient and timely disaster response. It is an utmost important to assess the surge capacity of the health care provider in one of the major districts in Sri Lanka, to obtain baseline data prior to initiate capacity development programs. This study provides important information about the existing surge capacity of healthcare institutions in Kurunegala district for the management of dengue outbreaks at a given time. Results of the present study can guide policy planners to initiate capacity building programs in line with the standards and guidelines and strategies. It can be used to develop education training programs and further research can be conducted to assess the response capacities of the healthcare workers in all public sector institutions for disaster management.

### Limitations of the study

There are different tools and methods for surge capacity assessment. However, there are limited studies on assessment of surge capacity. With the limited time and resources, this study was focused on basic assessment to have baseline data. There was no system to monitor equipment and their requirement. Therefore, the adequacy was measured subjectively. Equipment management systems need to be established in all healthcare institutions. Adequacy of healthcare staff was decided by the comparison with the cadre provision for the institutions. However, the cadre provisions for the institutions were estimated by the Ministry of Health in 2017. It needs to be updated at least annually according to the growing needs of the population. The study limited to one district out of 26 districts in Sri Lanka which partially reflects the situation in the country. Some of the results need to be further tested by further research choosing the other districts as well.

This study was limited to assess the surge capacity for disease outbreak management prior to COVID-19. Therefore, further research on disaster surge capacity needs to be conducted during this pandemic of COVID-19. All metrics for Science of Surge theory were not evaluated by this study due to non-availability of resources and time constraints.

## Declarations

### Ethics approval and consent to participate

Ethical approval was obtained from the ERC, Faculty of Medicine, University of Colombo, Sri Lanka (EC/18/134). Informed consent was obtained from all subjects. All methods were performed in accordance with the relevant guidelines and regulations.

### Consent for publication

Not applicable

### Availability of data and materials

The datasets used and/or analysed during the current study are available from the corresponding author on reasonable request.

### Funding for publication

This study did not receive any specific grant from funding agencies for publication.

### Authors’ contributions

Conceptualization of the study by RMNUR & CA. Implementation by RMNUR, CA, SM. Writing and Original draft preparation by RMNUR. Supervision, Review & Editing by CA, AB, MSDW, SM, YAA. All authors reviewed the manuscript.

## Data Availability

All data produced in the present study are available upon reasonable request to the authors

## Acknowledgements

North-western Province and Kurunegala District administrators and healthcare teams of Sri Lanka

## Authors’ information

### Principal Investigator PI]

RMNUR [Bachelor of Medicine and Bachelor of Surgery (MBBS); Doctor of Medicine (MD) in Public Health; Master of Science (MSc) in Public Health; Postgraduate Diploma in Health Sector Disaster Management, FRSPH (UK), Former Kurunegala District Disaster Focal Point

Senior Registrar in Community Medicine, Postgraduate Institute of Medicine, University of Colombo, Sri Lanka [Post-Doctoral Visiting Research Scholar, School of Public Health, University of Queensland, Australia]

### Main supervisor of the study

CA [MBBS, MSc in Community Medicine, MD in Community Medicine, PgDip Stat, BA, MA, MA, FCCP(SL)]

Senior Professor in Community Medicine, Department of Community Medicine, Faculty of Medicine, Ragama, University of Kelaniya, Sri Lanka

### Co-supervisor of the study

AB [MBBS, MSc in Community Medicine, MD in Community Medicine, PGDBA, MA], Professor, Consultant Community Physician, Faculty of Medicine, General Sir John Kotelawala Defence University, Ratmalana, Sri Lanka

### Other Co-authors

1. MSDW [MBBS, MSc Community Medicine, MD, MPH, MRSPH (UK)] Consultant Community Physician, Head / Family Health, Nutrition Communication & Behaviour Research Unit, Health Promotion Bureau, Ministry of Health, Sri Lanka
2. SM [MBBS, MD, MRCP(UK), FRCPL, FRCPE, FACP(USA), FCCP, MRCP Endocrinology & Diabetes (UK)], Consultant Physician in Internal Medicine, Teaching Hospital Kurunegala, Ministry of health, Sri Lanka.

## References

1. Hick, J. L., Koenig, K. L., Barbisch, D., & Bey, T. A. Surge capacity concepts for health care facilities: the CO-S-TR model for initial incident assessment. Disaster medicine and public health preparedness, 2008. 2 *Suppl 1*, S51–S57. 10.1097/DMP.0b013e31817fffe8

2. Hick, J. L., Hanfling, D., Burstein, J. L., DeAtley, C., Barbisch, D., Bogdan, G. M., & Cantrill, S. Health care facility and community strategies for patient care surge capacity. Annals of emergency medicine, 2004. 44(3), 253–261. 10.1016/j.annemergmed.2004.04.011

3. Koh, H. K., Shei, A. C., Bataringaya, J., Burstein, J., Biddinger, P. D., Crowther, M. S., Serino, R. A., Cohen, B. R., Nick, G. A., Leary, M. C., Judge, C. M., Campbell, P. H., Brinsfield, K. H., & Auerbach, J. Building community-based surge capacity through a public health and academic collaboration: the role of community health centers. 2006. Public health reports (Washington, D.C. : 1974), 121(2), 211–216. 10.1177/003335490612100219

4. Kaji, A., Koenig, K. L., & Bey, T. Surge capacity for healthcare systems: a conceptual framework. Academic emergency medicine : official journal of the Society for Academic Emergency Medicine, 2006. 13(11), 1157–1159. 10.1197/j.aem.2006.06.032

5. Adams, L. M. Exploring the Concept of Surge Capacity. (OJIN: The Online Journal of Issues in Nursing).2009. Retrieved from http://www.nursingworld.org/MainMenuCategories/ANAMarketplace/ANAPeriodicals/OJIN/TableofContents/Vol142009/No2May09/Articles-Previous-Topics/Surge-Capacity.html.

6. Barbisch, D. F., & Koenig, K. L. Understanding Surge Capacity: Essential Elements. Academic Emergency Medicine, 2006. 13(11), 1098–1102. Retrieved from, 10.1197/j.aem.2006.06.041

7. Phillips, S. Current status of surge research. Academic Emergency Medicine, 2006. 13, 1103–416.

8. Schultz, C.H. and Koenig, K.L. State of Research in High-consequence Hospital Surge Capacity. Academic Emergency Medicine, 2006. 13: 1153–1156. 10.1197/j.aem.2006.06.033

9. The American College of Emergency Physicians (ACEP). Health Care System Surge Capacity Recognition, Preparedness, and Response. 2007. Retrieved from https://www.acep.org/Clinical---Practice-Management/Health-Care-System-Surge-Capacity-Recognition,-Preparedness,-and-Response/#sm.0001x5vr14li2dovu0426pyhn0kdk.

10. WHO. Disease outbreaks- WHO.2017. Retrieved from http://www.who.int/topics/disease_outbreaks/en/.

11. WHO. Dengue control: Monitoring and evaluation of programmes. 2018. Retrieved from http://www.who.int/denguecontrol/monitoring/behaviour/en/.

12. The National Academy of Sciences. Regional Disaster Response Coordination to Support Health Outcomes: Summary of a Workshop Series.2015. doi: Bookshelf ID: NBK311257

13. Tissera H, Pannila-Hetti N, Samaraweera P, Weeraman J, Palihawadana P, Amarasinghe A. Sustainable dengue prevention and control through a comprehensive integrated approach: the Sri Lankan perspective. WHO South-East Asia Journal of Public Health | September 2016 | 5 (2), 106–112. Retrieved from http://www.searo.who.int/publications/journals/seajph/issues/seajph2016v5n2p106.pdf?ua=.

14. Hanfling, D. Equipment, Supplies, and Pharmaceuticals: How Much Might It Cost to Achieve Basic Surge Capacity? Academic Emergency Medicine, 2006. 13(11), 1232–1237. 10.1197/j.aem.2006.03.567

15. Davis, A. and Lemma, T. Capacity Development : A UNDP Primer. 2009. Retrieved from www.undp.org/capacity

16. Runge-Ranzinger S, Kroeger A, Olliaro P, McCall PJ, Sánchez Tejeda G, Lloyd LS, et al.. Dengue Contingency Planning: From Research to Policy and Practice. PLoS Negl Trop Dis 2016.10(9): e0004916. doi:10.1371/journal.pntd.0004916, https://www.ncbi.nlm.nih.gov/pmc/articles/PMC5031449/pdf/pntd.0004916.pdf

17. WHO/ EHA. Disasters and emergencies, definitions-Training package. 2002. http://apps.who.int/disasters/repo/7656.pdf.

18. Handler, J. A., Gillam, M., Kirsch, T. D., & Feied, C. F. Metrics in the Science of Surge. Academic Emergency Medicine, 2006. 13(11), 1173–1178. 10.1197/j.aem.2006.07.006

19. Zhiheng Z., Caixia, W., Jiaji, W., Huajie, Y., Chao, W., & Wannian, L. The knowledge, attitude, and behavior about public health emergencies and the response capacity of primary care medical staffs of Guangdong Province, China. BMC Health Services Research 2012, 12:338, 1–9. doi: 10.1186/1472-6963-12-338.

20. Kumar, A., & Weibley, E. Disaster management and physician preparedness. Southern medical journal, 2013. 106(1), 17–20. 10.1097/SMJ.0b013e3827c5c5b

21. Uddin, S. G., Barnett, D. J., Parker, C. L., Links, J. M., & Alexander, M. Emergency preparedness: addressing a residency training gap. Academic Medicine: Journal of the Association of American Medical Colleges, 2008. 83(3), 298–304. 10.1097/ACM.0b013e3181637edc

22. Srichan P, Niyom SL, Pacheun O, Iamsirithawon S, Chatchen S, Jones C, White LJ, Pan-Ngum W. Addressing challenges faced by insecticide spraying for the control of dengue fever in Bangkok, Thailand: a qualitative approach. Int Health. 2018 Sep 1;10(5):349–355. doi: 10.1093/inthealth/ihy038. PMID: 29912451; PMCID: PMC6104709.

23. Dasanayaka S. Performance of Health Care Equipments in the Public Sector Hospitals in the Eye of Good Governance, A Case Study Based on the Sri Lankan Public Sector Hospitals. 2011. Retrieved from https://www.researchgate.net/publication/228760950_of_Health_Care_Equipments_in_the_Public_Sector_Hospitals_in_the_Eye_of_Good_Governance_A_Case_Study_Based_on_the_Sri_Lankan_Public_Sector_Hospitals

24. WHO. The World Health Report. 2006. Working Together for Health, Geneva.

25. Cantrill, S., Bonnett, C., Hanfling, D., & Pons, P. Alternative care sites. In Phillips, S.J., Knebel, A. (eds.) (2007). Mass medical care with scarce resources: A community planning guide. (AHRQ Publication No. 07-0001, 2007. pp.75-99). Rockville, MD: Agency for Healthcare Research and Quality.

26. Christian, M.D., Devereaux, A.V., Dichter, J.R., Geiling, J.A., & Rubinson, L. (2008). Definitive care for the critically ill during a disaster: Current capabilities and limitations. CHEST, 133(*Supplement*), 8S–17S.

27. Assistant Secretary for Preparedness and Response (ASPR). What is Medical Surge?. 2012. Retrieved from https://www.phe.gov/Preparedness/planning/mscc/handbook/chapter1/Pages/whatismedicalsurge.aspx.

28. Agency for Healthcare Research and Quality (AHRQ). Annual Patient Safety and Health Information Technology Conference:2005. Retrieved from; https://digital.ahrq.gov/2005-annual-conference

29. Jayawardena A. Hospital Equipment Management in District Base Hospitals in Kalutara District in Sri Lanka. 2017. Retrieved from, https://www.researchgate.net/publication/326440810_Hospital_Equipment_Management_in_District_Base_Hospitals_in_Kalutara_District_in_Sri_Lanka

30. Sumathipala, A., Siribaddana, S., & Perera, C. Management of dead bodies as a component of psychosocial interventions after the tsunami: A view from Sri Lanka. International Review of Psychiatry, 2006.18, 249–257. Retrieved from; https://www.semanticscholar.org/paper/Management-of-dead-bodies-as-a-component-of-after-A-Sumathipala-Siribaddana/8b928941fb9b0e528e237447857ea04fb0cd9ca6

31. DPRD. Strategic Plan for Health Sector Disaster/Emergency Preparedness. Sri Lanka : Ministry of Health. 2011

32. Al-Ali, N.M. & Ibaid, A.H.A. Health-care providers’ perception of knowledge, skills and preparedness for disaster management in primary health-care centers in Jordan. Eastern Mediterranean Health Journal, 2015. 21(10), 713–721.

33. Rajapaksha, N.U., Abeysena, C., Balasuriya, A. et al. Incidence management system of the healthcare institutions for disaster management in Sri Lanka. BMC Emerg Med 23, 6 (2023). 10.1186/s12873-023-00777-y

